# Reward enhances motor learning in acute stroke patients by reducing motor noise

**DOI:** 10.1101/2024.04.03.24305243

**Authors:** Theresa Paul, Valerie M. Wiemer, Jonas G. Nix, Finn M. Lehnberg, Scott T. Grafton, Gereon R. Fink, Lukas J. Volz

## Abstract

The majority of motor recovery occurs within the first weeks after stroke and partially relies on similar mechanisms as motor learning in the healthy brain. Given that motor learning can result from both error- and reinforcement-based mechanisms, we investigated whether complementing error-based adaptation with reinforcement feedback enhances motor learning early after stroke. Here, we show for the first time that acute stroke patients exhibit successful error-based visuomotor adaptation with their paretic hand. Reward and punishment feedback exerted opposite modulatory effects on motor adaptation: while performance-dependent punishment feedback hampered adaptation, reward enhanced both initial learning and retention. Mechanistically, reward provided a complementary teaching signal when sensory prediction errors were compromised by sensorimotor deficits resulting in a systematic reduction of motor noise. Our results emphasize that combining reward feedback with motor adaptation facilitates relearning of motor control after stroke and may thus be critical to enhance motor recovery in future therapeutic settings.

## Main

Motor impairments after stroke are exceedingly common and represent a leading cause of reduced quality of life and loss of independence.^1,2^ While motor recovery is thought to arise from a relearning of motor control akin to motor learning in the healthy brain,^3^ our understanding of motor learning after stroke remains rudimentary.^4^ Closing this gap seems particularly relevant as the acquisition and adaptation of motor control policies can arise from various mechanisms including error- and reinforcement-based motor learning,^5–7^ potentially providing the opportunity to compensate for stroke-induced impairment in a particular learning domain by an increased reliance on alternative learning mechanisms.^8^

Error-based motor learning can be studied via *motor adaptation*, which describes how the motor system accounts for perturbations such as sudden changes in biomechanical properties or external conditions.^9,10^ In the healthy brain, error-based motor adaptation learning can be selectively modulated by *reinforcement feedback*. A seminal visuomotor adaptation study by Galea and colleagues reported dissociable effects of reward and punishment.^11^ Whereas punishment accelerated adaptation, reward enhanced retention, i.e., how well the newly learned motor control policy was internalized and recalled. While the exact underlying mechanisms remain unknown, the beneficial effect of reward on adaptation has been attributed to a reduction in motor noise.^12^

After stroke, such adaptation paradigms seem particularly well-suited to investigate the relearning of lost upper-limb motor function as they allow to assess how the motor system updates its control policies to overcome movement errors caused by a sudden perturbation. Of note, several studies have reported that chronic stroke patients possess the ability to perform motor adaptation using their paretic hand,^13–17^ and deficient visuomotor adaptation has been linked to the severity of motor impairment.^15^ Regarding the modulatory effect of reinforcement, faster learning rates have been found in response to both punishment and reward in chronic stroke patients, yet only reward facilitated retention.^17^ Importantly, studies investigating motor adaptation learning and the modulatory influence of reinforcement feedback on adaptation in acute patients are missing to date.^18^ Given that the vast majority of motor recovery occurs during a phase of elevated plasticity within the first weeks after stroke,^19^ it seems critical to further our understanding of how error- and reinforcement-based motor learning contribute to and shape early motor recovery.

Therefore, we aimed to answer two major questions: (i) Can acute stroke patients perform motor adaptation with their paretic hand? (ii) If so, can reinforcement feedback enhance motor adaptation and retention in acute stroke patients? In line with previous findings,^11,20^ we hypothesized that punishment might speed up initial motor learning but anticipated no impact on retention. In contrast, we expected a beneficial effect of reward on both adaptation and retention in line with previous studies in healthy participants and chronic stroke patients.^11,17^ From a mechanistic perspective, we anticipated beneficial effects of reward to partially derive from a reduction in motor noise as suggested by previous results in healthy subjects.^12^

Our results showed that acute stroke patients exhibited adaptation of motor control with their paretic hand and successfully retained updated control policies over time. While punishment feedback had an unexpected detrimental effect on motor performance, reward enhanced both adaptation learning and retention by reducing disruptive motor noise. Mechanistically, this systematic reduction in motor noise may result from the teaching signal of performance-dependent reward feedback, which might help to guide motor learning when sensory prediction errors are compromised by the stroke-induced sensorimotor deficit. From a clinical perspective, strategically enhancing motor adaptation during a period of heightened plasticity early after stroke via reward feedback holds seminal implications for advancing rehabilitation and motor recovery.

## Results

### Motor adaptation in acute stroke patients

Acute stroke patients performed a joystick-based visuomotor adaptation task with their paretic hand consisting of center-out reaching movements (see Figure 1 for an overview). In three sessions conducted on subsequent days, a distinct reinforcement condition (neutral, reward, or punishment) was introduced in a counter-balanced order across participants. During each session, patients first underwent a learning phase during which two adaptation blocks featuring a suddenly introduced visuomotor rotation (*adaptation 1* and *adaptation 2*) were interleaved with blocks of non-rotated trials (*baseline* and *de-adaptation*). After a 30-minute break, we probed for retention effects with a block of rotated trials (*retention*) followed by an unrotated washout phase (*washout*).

**Figure 1:**
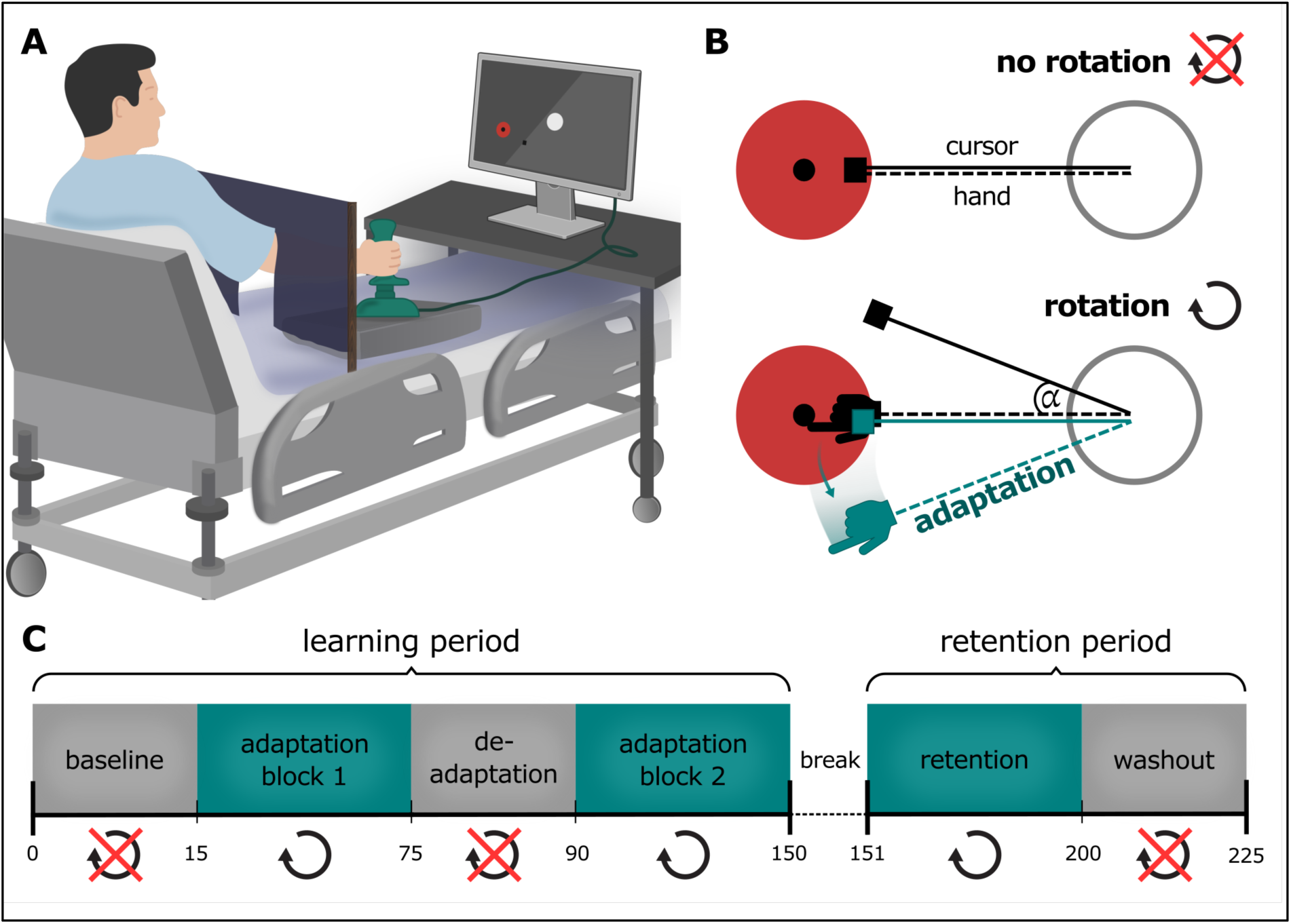
Visuomotor adaptation task. (A) Using a joystick, patients performed center-out reaching movements with their paretic hand in a bedside manner. Once the joystick was centered, the outside target turned red, signaling the start of the movement. During the movement, participants continuously saw the cursor on the screen. A trial ended as soon as the cursor reached the outside target. The light tug of the joystick’s spring then facilitated a smooth and effortless movement back to the center. Of note, patients were unable to see their arm or hand, which were hidden by a curtain during task performance. (B) In the “no rotation” condition, veridical cursor feedback was provided. In other words, the hand movement matched the visual feedback on the screen. Conversely, in the “rotation” condition, the cursor feedback was rotated by a fixed rotation angle α, so that the visual cursor information deviated from the actual hand movement (black line). Patients thus had to compensate for the rotation (green line) to achieve a straight line from center to target. (C) The experiment consisted of a learning and retention period. Each of these periods contained blocks with and without a visuomotor rotation. Notably, while reward, punishment, or neutral feedback were provided in the learning period, the retention period was free of reinforcement feedback in all sessions.

Motor performance was quantified as *movement error*, computed as the deviation from the ideal trajectory from center to target^21^ (see Materials and Methods for a detailed description). For model-free analyses, these trial-wise movement errors were statistically compared, while model-based analyses derived learning rates based on movement error data using a single-rate state-space model in line with previous research.^11^

### Successful adaptation and savings

We assessed established hallmarks of motor adaptation learning in the present sample of acute stroke patients. A model-free comparison of early and late trials in adaptation block 1 across all reinforcement conditions (average movement error of the first 15 versus the last 15 trials) indicated that patients exhibited successful motor adaptation (t(65) = 7.97, p < 0.001; Figure 2). Moreover, comparing average performance across the first 15 trials of adaptation blocks 1 and 2 revealed *savings*. In other words, adaptation was faster (t(65) = 3.02, p = 0.004) when patients were exposed to the same rotation for a second time. In line with the model-free results, *savings* were also observed in model-based analyses with the second adaptation block yielding faster learning rates than the first adaptation block (t(65) = −4.15, p < 0.001). Notably, we found a significant negative correlation between initial learning rates derived from adaptation block 1 and average movement errors of the retention block (r = −0.50, p < 0.001). Thus, faster initial learning was associated with improved performance after training, consistent with the notion that successful early adaptation enhances the retention of updated motor control policies.

**Figure 2:**
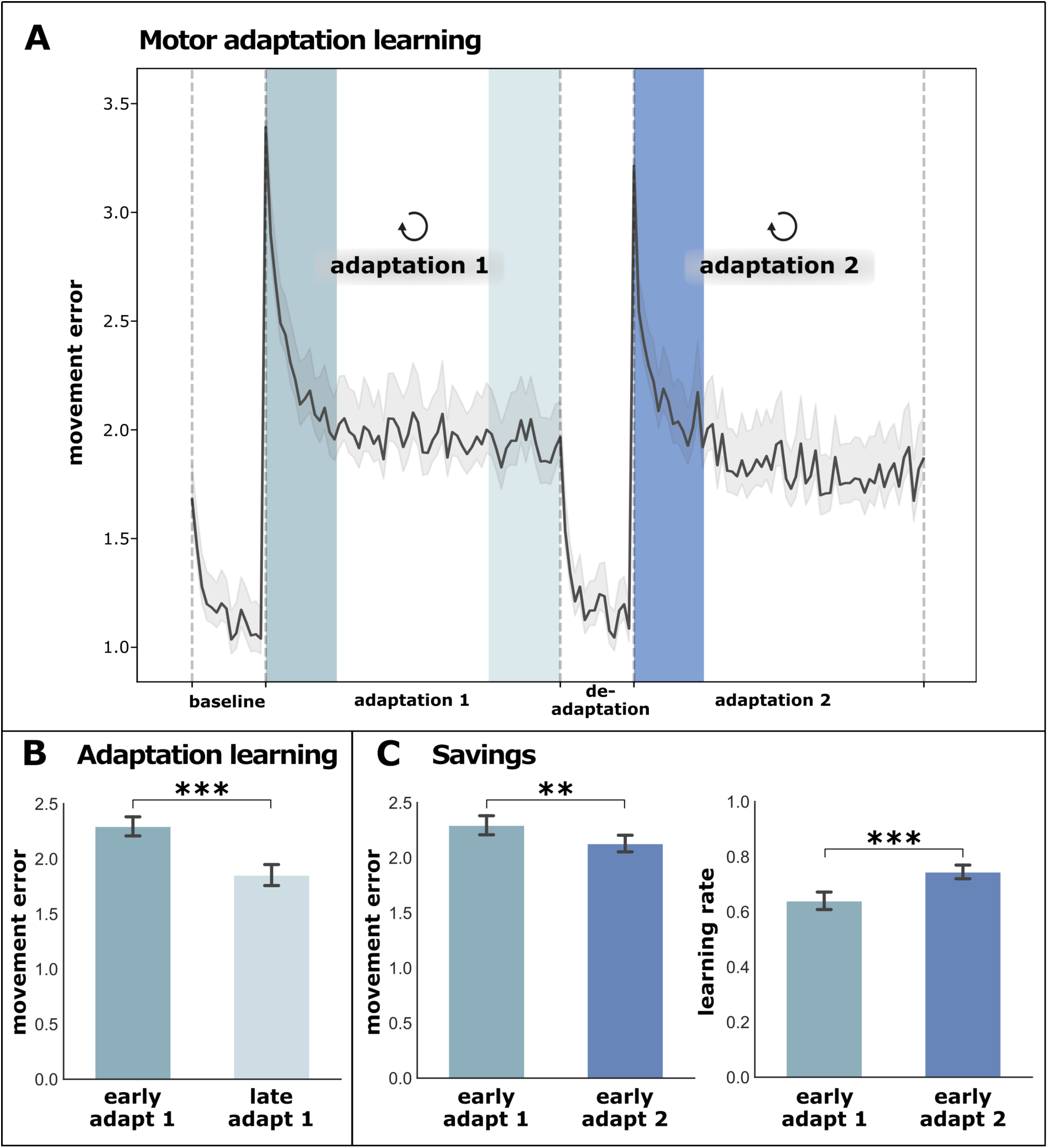
Initial motor adaptation learning. (A) Movement error is denoted over time across patients, with lower movement errors indicating better performance. (B) Comparing performance during the first and last 15 trials of the first adaptation block showed that patients’ performance improved significantly. This finding indicates that patients were able to adapt their movements to compensate for the rotation introduced at the beginning of this block. (C) When comparing patients’ performance during the first 15 trials of adaptation blocks 1 and 2, we observed significantly better performance in adaptation block 2 when the visuomotor rotation was introduced for a second time. Thus, patients adapted faster when recalling the new motor control policy, thereby showing *savings*. Faster adaptation upon reintroduction of the visuomotor rotation was confirmed by significantly higher (i.e., faster) learning rates in adaptation block 2 derived from model-based analyses.

### Reward enhances motor learning after stroke

Next, we tested whether reward and punishment feedback affected motor adaptation and retention by computing repeated measures ANOVAs with the within-subject factor reinforcement condition (neutral, reward, punishment) for adaptation block 1 as well as the retention block using movement errors (model-free analysis) or learning rates (model-based analysis) as dependent variables.

Model-free analyses revealed a main effect of reinforcement condition during adaptation block 1 (F(2,42) = 3.74, p = 0.032, generalized η^2^ = 0.05; Figure 3). FDR-corrected paired-sample post-hoc t-tests yielded a significant difference between reward and punishment (t(21) = −2.83, p = 0.030). While there was no difference between conditions in adaptation block 2 (F(2,42) = 1.30, p = 0.282), movement errors significantly differed between conditions during the retention block (F(2,42) = 6.22, p = 0.004, generalized η^2^ = 0.06). Specifically, participants performed significantly better in the reward condition than in the neutral (t(21) = 2.47, p = 0.033) or punishment (t(21) = −2.95, p = 0.023) conditions. Of note, the beneficial effect of reward on movement errors extended into the washout phase (F(2,42) = 3.66, p = 0.049, generalized η^2^ = 0.04) with a significant difference between reward and punishment (t(21) = −2.61, p = 0.049). To rule out the possibility that reward simply enhanced overall motor performance rather than learning, we tested for reinforcement effects during the baseline block, which did not yield any significant differences (F(2,42) = 0.09, p = 0.869).

**Figure 3:**
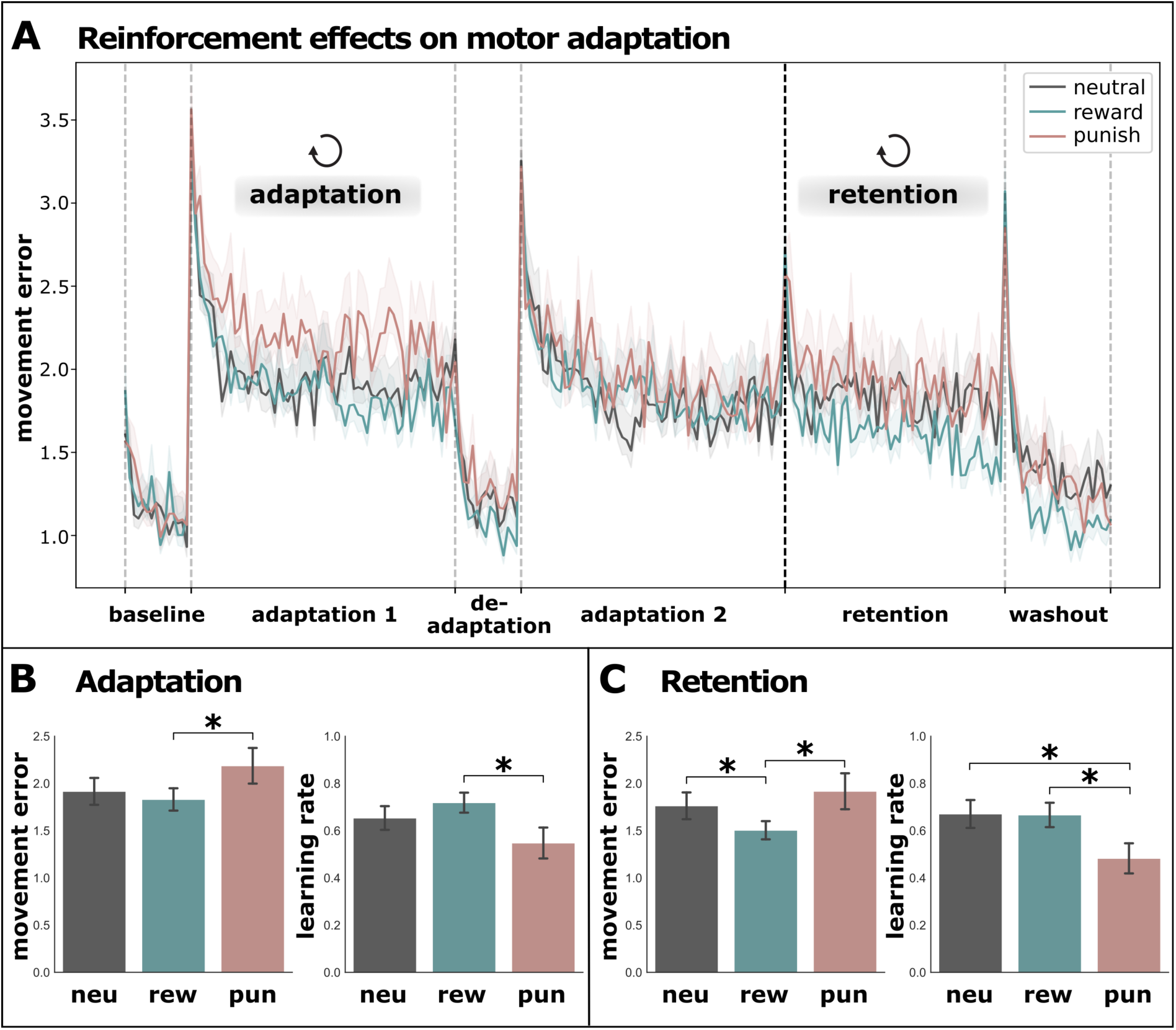
Reinforcement effects on motor adaptation. (A) Trial-wise movement errors per condition are depicted over time with lower movement errors indicating better performance. (B) In the first adaptation block, patients performed better in the reward than in the punishment condition, reflected in both the model-free analysis (lower movement errors) and model-based analysis (higher learning rates). Thus, patients not only showed poorer performance when receiving performance-dependent punishment but also exhibited slower adaptation to the visuomotor rotation. (C) In the retention phase, patients performed best when the prior learning period was reinforced by performance-dependent reward. Of note, comparing learning rates showed that patients adapted more slowly with prior punishment compared to previous neutral or reward feedback. Thus, reward feedback during the learning period improved retention of the novel motor control policy. Moreover, while neutral or punishment feedback led to a stable performance level after a 30-minute break (comparing adaptation block 2 and retention), significant improvements after the break were observed in the reward condition. *adapt = adaptation, neu = neutral, rew = reward, pun = punishment*

Comparing the learning rates derived from the model-based approach, we observed highly similar results. We found a main effect of reinforcement in adaptation block 1 (F(2,42) = 3.35, p = 0.045, generalized η^2^ = 0.08) with patients learning faster in the reward than in the punishment condition (t(21) = 2.70, p = 0.041). While no difference in learning rates was observed for adaptation block 2 (F(2,42) = 2.32, p = 0.111) in line with model-free results, learning rates significantly differed between conditions during retention (F(2,42) = 4.62, p = 0.015, generalized η^2^ = 0.10). Of note, patients exhibited slower learning rates in the retention period when they received punishment during the preceding learning phase compared to neutral feedback (t(21) = 2.66, p = 0.044) or reward (t(21) = 2.33, p = 0.045), despite the fact that no further reinforcement was presented in the retention phase in any condition.

When testing for retention effects, we observed a clear dissociation between reward and punishment. Punishment and neutral feedback during initial learning led to successful retention, as indicated by comparable movement errors during adaptation block 2 and retention (neutral: t(21) = −0.10, p = 0.921; punishment: t(21) = −0.01, p = 0.991). Conversely, reward feedback during initial learning resulted in performance gains, reflected by lower movement errors during the retention period than during adaptation block 2 (t(21) = 3.41, p = 0.003; average movement error adaptation 2 = 1.78, average movement error retention = 1.50).

### Speed-accuracy trade-off and motor learning

Motor performance can be differentiated with regard to two fundamental features: speed and accuracy.^12^ When improvements in one domain occur at the cost of the other, the change in performance cannot be considered a true improvement but rather reflects a shift in the speed-accuracy trade-off. Thus, genuine learning can be assumed to occur when (i) both domains exhibit improvements, or (ii) one domain shows improvements while the other remains constant. To independently assess speed and accuracy, we quantified movement duration and movement inaccuracy (see Materials and Methods for a detailed description). Model-free analyses yielded a significant improvement of both movement duration (t(65) = 5.48, p < 0.001) and movement inaccuracy (t(65) = 7.82, p < 0.001) for adaptation block 1 when comparing the first to the last 15 trials (Figure 4). In line with the movement error results, we also observed savings for inaccuracy t(65) = 3.22, p = 0.002) and a statistical trend towards savings for movement duration (t(65) = 1.86, p = 0.067).

**Figure 4:**
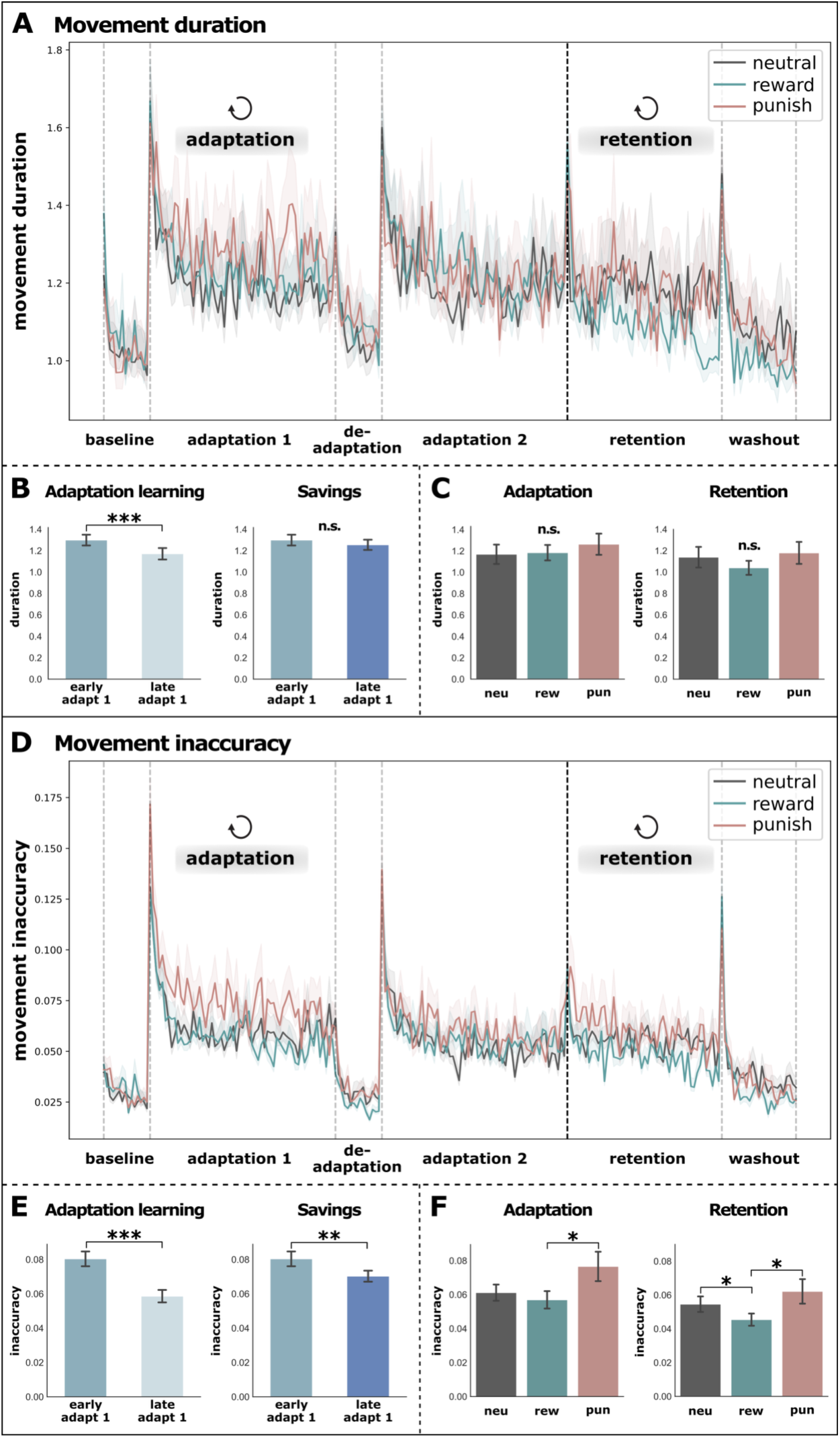
Speed-accuracy trade-off. To test whether patients exhibited genuine motor learning rather than a mere shift along the speed-accuracy trade-off continuum, both domains were assessed independently. We therefore recomputed all analyses conducted for the movement error using *movement duration* and *movement inaccuracy*. (A&D) Overall, similar improvements in performance were observed for both measures compared to movement errors. (B&E) In particular, significant improvements over the course of the first adaptation block were observed for both movement inaccuracy and movement duration, indicating that patients simultaneously became faster and more accurate. Moreover, significant savings were observed for movement inaccuracy. (C) While movement duration did not differ across reinforcement conditions, (F) similar reinforcement effects emerged for movement inaccuracy compared to movement error for the adaptation and retention phases. Thus, these results highlight a beneficial effect of reward feedback on movement accuracy while speed remained constant. Overall, our findings emphasize that patients exhibited actual motor learning rather than a shift in the speed-accuracy trade-off with genuine improvements achieved through reward feedback. *adapt = adaptation, neu = neutral, rew = reward, pun = punishment*

When repeating the analyses of reinforcement effects using movement inaccuracy rather than movement error as the dependent variable in a repeated-measures ANOVA, we observed highly similar results. For adaptation 1, movement inaccuracy (F(2,42)= 4.63, p = 0.015, generalized η^2^ = 0.08) showed a main effect of reinforcement with significantly better adaptation performance during reward than during punishment (t(21) = −2.74, p = 0.037). For the retention phase, we also observed a main effect of reinforcement for inaccuracy (F(2,42) = 4.92, p = 0.012, generalized η^2^ = 0.07), with reward leading to better retention compared to neutral (t(21) = 2.66, p = 0.034) or punishment feedback (t(21) = −2.46, p = 0.034). For movement duration, we did not observe effects of reinforcement in the first adaptation block (F(2,42) = 1.78, p = 0.180) but for the retention phase (F(2,42) = 3.80, p = 0.030, generalized η^2^ = 0.02). However, post-hoc t-tests did not show any significant differences between conditions after FDR-correction (all p > 0.05). This suggests that movement duration remained constant across reinforcement conditions, indicating that changes in response to reinforcement feedback did not emerge at the cost of movement speed.

In summary, our data highlight that acute stroke patients simultaneously improved in both movement inaccuracy and movement duration, emphasizing that patients indeed exhibited motor learning. Moreover, the effects of reward and punishment were only observed for changes in accuracy, while movement speed was comparable across reinforcement conditions. Hence, our results are consistent with the notion that reward led to an actual improvement in motor adaptation rather than a shift in the speed-accuracy trade-off.

### Reward reduces motor noise

To test whether performance-dependent reinforcement can serve as a teaching signal to guide motor learning when sensory prediction errors are compromised early after stroke, we assessed the immediate trial-wise after-effects of reinforcement. To do so, we compared motor noise and movement inaccuracy in trials immediately following a reinforced versus non-reinforced trial within the first adaptation block (Figure 5). In line with previous research, motor noise was defined as the standard deviation of movement inaccuracy.^22^ Our results showed that both motor noise (t(65) = 3.36, p = 0.001) and movement inaccuracy (t(65) = 2.00, p = 0.029) decreased in trials following rewarded compared to non-rewarded trials. Thus, motor performance was better after rewarded than non-rewarded trials in the reward condition. Conversely, in the punishment condition, motor noise (t(65) = 1.72, p = 0.050) and movement inaccuracy (t(65) = 4.21, p < 0.001) increased following punished trials compared to trials eliciting no punishment. Thus, punishment feedback led to higher motor noise and poorer motor performance in subsequent trials.

**Figure 5:**
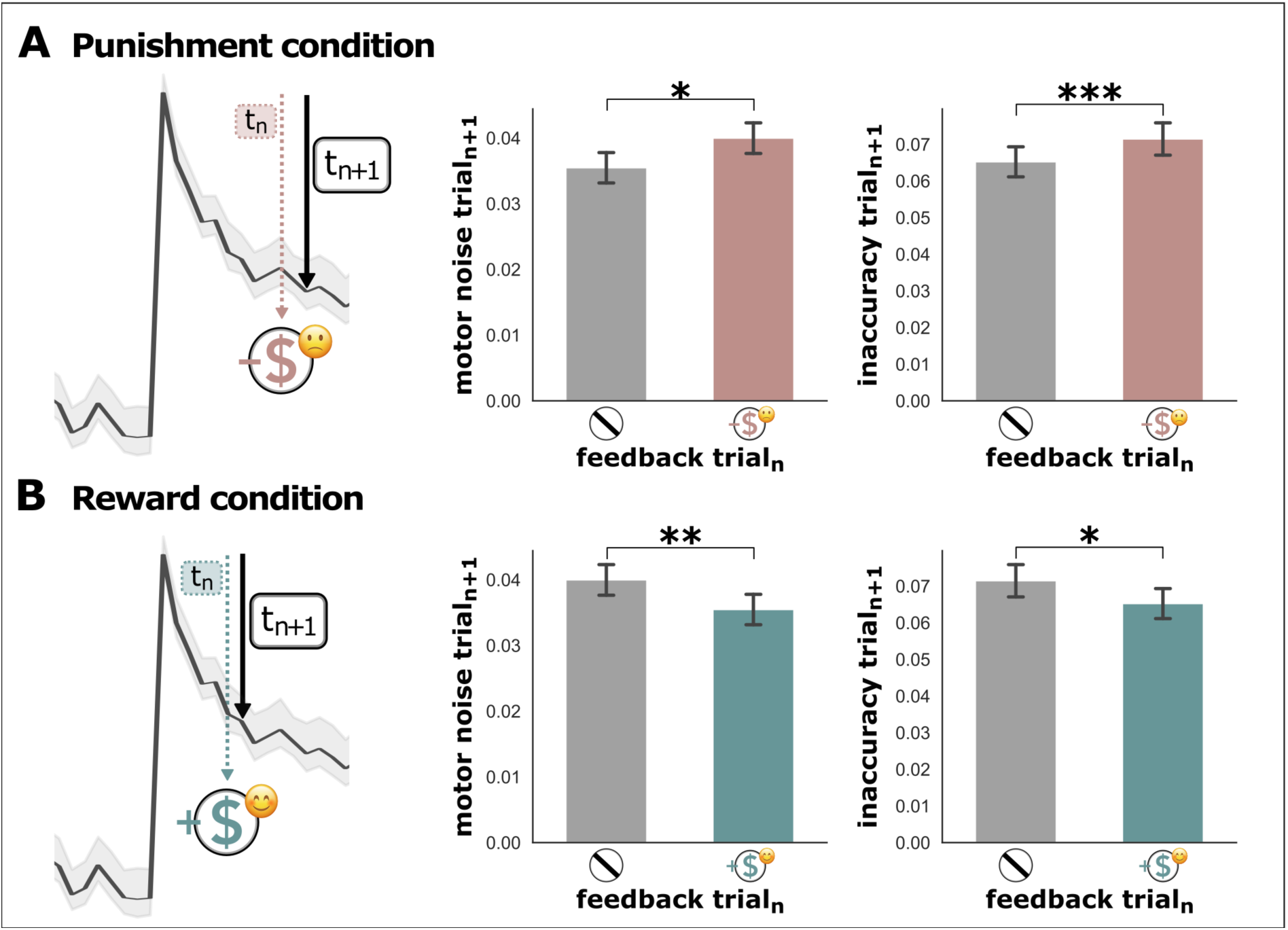
Motor noise and movement inaccuracy after reinforced trials. To assess the immediate effect of performance-dependent reward and punishment feedback on subsequent motor execution, we analyzed motor noise (i.e., the standard deviation of movement inaccuracy) and movement inaccuracy in trials (t_n+1_) immediately following reinforcement feedback in the previous trial (t_n_) within the first adaptation block. (A) Punishment condition: When comparing trials t_n+1_ following punishment feedback with trials t_n+1_ following no feedback in the same adaptation block, we observed significantly higher levels of motor noise and movement inaccuracy in the post-punishment trials. (B) Reward condition: Conversely, trials t_n+1_ after reward feedback featured lower levels of motor noise as well as lower levels of movement inaccuracy than trials t_n+1_ following no feedback. Thus, our results highlight a differential immediate effect of reward and punishment feedback on motor noise and performance: While punishment increased motor noise and hampered motor performance in the subsequent trial, reward feedback led to an immediate improvement in motor noise and performance.

Next, we tested whether the beneficial effect of reward on subsequent trials led to an overall reduction in motor noise. We therefore quantified blockwise motor noise as the standard deviation of movement inaccuracy across adaptation block 1. Comparing motor noise across reinforcement conditions using a repeated measures ANOVA, we observed a main effect of reinforcement in motor noise for adaptation 1 (F(2,42) = 5.72, p = 0.006, generalized η^2^ = 0.08), with lower levels of motor noise during reward than punishment (t(21) = −3.75, p = 0.004) and a trend towards lower levels of motor noise during the neutral than punishment condition (t(21) = −2.05, p = 0.08). Thus, while reward feedback reduced motor noise, punishment appeared to introduce additional motor noise to the system.

To assess whether the level of motor noise may influence the subsequent retention of novel motor control policies, we probed for an association between motor noise during the learning phase and movement errors during retention. Of note, motor noise during adaptation 1 was significantly associated with performance in the retention phase. In particular, lower motor noise was associated with lower movement errors in the retention (r = 0.50, p < 0.001). In line with these findings, we also observed a negative correlation between initial learning rates in adaptation 1 and motor noise, emphasizing the negative impact of motor noise on motor adaptation (r = −0.31, p = 0.011).

In summary, these findings highlight that the beneficial effect of reward on motor adaptation may be mediated by a systematic reduction in motor noise. From a mechanistic perspective, the performance-dependent reward may serve as a teaching signal instructing the motor system to use a highly similar control policy in the upcoming trial, thereby reducing noise across trials. Consequently, performance-dependent reward feedback may guide adaptation when error-based signals are compromised by the sensorimotor deficit in the acute phase after stroke.

## Discussion

### Successful motor adaptation in acute stroke

While recovery of stroke-induced motor impairment has frequently been discussed to rely on similar mechanisms as motor learning in the healthy brain,^3,5^ our understanding of motor adaptation early after stroke remains highly limited.^23^ Regarding reaching movements performed with the paretic arm, successful adaptation and retention thereof have been shown in chronic stroke patients.^17^ Moreover, inter-individual deficits in visuomotor adaptation have recently been discussed to be associated with the level of clinical impairment across patients.^15^ Notably, the ability to successfully exhibit motor adaptation may indicate the motor system’s potential to flexibly compensate for the effect of the stroke lesion, which can hence be considered as a special case of motor adaptation. Here, we extend our current knowledge by showing, for the first time, that acute stroke patients can indeed perform motor adaptation with their paretic hand. More specifically, we replicated hallmarks of adaptation learning typically observed in healthy subjects, i.e., successful adaptation^24^ and savings across blocks.^25,26^ Of note, further disentangling the contributions of improvements in speed and accuracy indicated that patients exhibited improvements in both domains, highlighting genuine motor learning rather than a mere shift along the speed-accuracy continuum.^12^ Thus, our data show that adaptation mechanisms support relearning of motor control policies and thereby facilitate motor recovery after stroke.

### Punishment hinders early motor learning after stroke

Motor adaptation is driven by sensory prediction errors and heavily relies on a cerebellar forward model, which integrates predicted and actual sensorimotor states.^27–31^ In healthy subjects, punishment but not reward feedback has been shown to increase initial learning rates during motor adaptation, potentially via an increased sensitivity to sensory prediction errors.^11,20^ Notably, reinforcement feedback may be especially suited to guide motor learning when the quality of sensory feedback is low.^6,7^ In this case, a decreased informational value of sensory prediction errors may be compensated via an increased reliance on reinforcement signals. In line with this notion, both punishment and reward have been shown to enhance initial adaptation in chronic stroke patients.^17^ In contrast, we observed that punishment negatively affected patients’ motor performance during the initial learning phase compared to reward (Figure 3). An explanation for this detrimental effect of punishment may arise from a decrease in its informational value. Findings in healthy subjects highlight that for punishment feedback to be effective it needs to be informative, i.e., coupled to motor performance in a meaningful way.^11^ Given that the forward model is still undergoing recalibration to account for the lesion-induced impairment early after stroke,^10^ the repertoire of alternative available motor control policies is limited. In other words, punishment feedback imposes a challenge for the stroke-afflicted motor system: while punishment feedback clearly signals a switch to a different motor control policy in the next trial, it does not help to specify which subset of motor control policies might be preferable. Thus, punishment feedback does not provide information about how to update the forward model. As the motor system is in a state of reorganization, it is unable to select a more appropriate control strategy from its existing repertoire, which may render punishment feedback largely useless early after stroke.

### Reward feedback reduces motor noise

In contrast to punishment, performance-dependent reward feedback may indeed serve as a meaningful teaching signal helping to guide motor adaptation early after stroke. As motor execution is an ill-defined problem, i.e., a task with more than one solution with regard to the interplay of motor synergies, it is paramount for consistent performance to find an (almost) optimal solution that minimizes the influence of intrinsic motor noise and maximizes error tolerance.^32^ In order to avoid getting stuck in a local maximum, participants might explore the motor space in the beginning before settling on a strategy which exploits ideal motor synergies that yield minimal variability by being robust to intrinsic motor noise.^33^ Mechanistically, reward feedback might facilitate finding an optimal solution as it signals that the previous motor control policy led to successful task execution. In particular, performance-dependent reward may help inform the forward model that the previous motor control policy should be repeated. Indeed, we found that patients showed reduced motor variability and improved accuracy in trials immediately following rewarded trials (Figure 5). Conversely, motor noise and movement inaccuracy were higher in trials following punishment feedback. Moreover, reward led to lower levels of overall motor noise during the first adaptation block compared to punishment. This nicely aligns with earlier findings by Manohar and colleagues who reported that reward resulted in increased speed and accuracy by reducing internal motor noise. Within their cost-of-control framework, they posited that reward facilitated motor learning by “paying the cost of noise reduction”.^12^ Of note, one might argue that besides detrimental motor noise hindering the motor system to adequately learn from sensory prediction errors,^34^ variability in motor control may reflect meaningful exploration during the search for an optimal solution.^22,35^ However, in acute stroke patients who suffer from unreliable sensory prediction errors due to acute sensorimotor deficits, the variability in motor execution attributable to internal motor noise can be expected to be higher than a potential gain in informational value achieved via motor exploration.^36^ In line with the notion that motor variability early after stroke may primarily reflect increased levels of motor execution noise,^34^ we observed a negative correlation between learning rates and motor variability.

At first glance, our findings and interpretation might seem at odds with a recent account of impaired reinforcement-based motor learning in acute stroke patients.^37^ However, the study assessed reinforcement learning in isolation from error-based learning rather than combining both learning mechanisms in a single task. It therefore seems likely that improvements in motor learning strongly depend on the simultaneous engagement in error- and reinforcement-based learning. This notion is corroborated by Nikooyan and colleagues who showed that the combination of reward and sensory feedback improved motor adaptation in healthy subjects compared to error- or reinforcement-based learning alone.^38^ Thus, reward signals might help the damaged motor system to better interpret sensory prediction errors, which may in turn reduce motor noise and facilitate motor learning (see Figure 6 for a conceptual framework).

**Figure 6:**
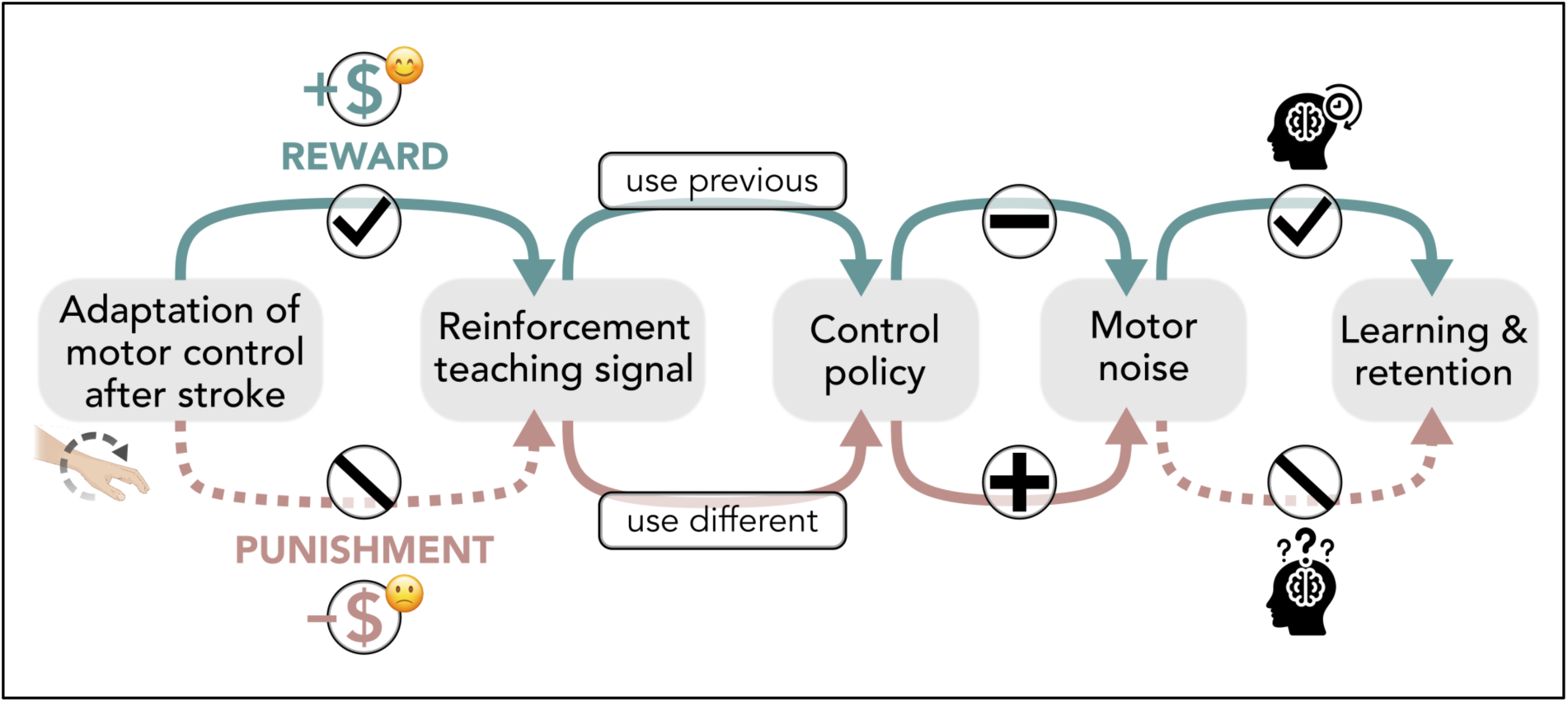
Conceptual framework of error- and reinforcement-based motor adaptation after stroke. We combined a visuomotor adaptation paradigm based on sensory prediction error learning mechanisms with performance-dependent reward or punishment feedback. Our results suggest that reward but not punishment feedback facilitates motor adaptation when sensory prediction errors are compromised due to acute sensorimotor deficits. The reward feedback may serve as a teaching signal informing the motor system to re-use the previous successful motor control policy. This teaching signal may lead to a reduction of motor noise, thereby enhancing learning and retention. Conversely, when a poorly executed trial is being punished, the motor system may be incentivized to change its control policy; however, no information is provided on how to adapt it. As a result, subsequent motor synergies are highly variable, increasing motor noise and ultimately hindering motor adaptation and retention.

### Clinical implications

Given the rising number of stroke patients,^2^ there is an urgent need for improved rehabilitation strategies that target the relearning of motor control. In this context, positive effects on retention are most critical as they may amplify the lasting effects of rehabilitative training. Importantly, performance-dependent reward feedback during the learning period led to better retention even when reinforcement feedback was no longer present (Figure 3). Given the observed negative effects of punishment, our findings discourage the application of negative feedback in a therapeutic setting, while providing the first evidence for a positive effect of reward on error-based motor learning in acute stroke patients. This nicely aligns with previous reports emphasizing the beneficial effect of reward on motor retention in healthy individuals^11,39–41^ and chronic stroke patients.^17^ Of note, the positive effect of reward feedback on retention has been shown to persist for up to 30 days after motor learning in healthy subjects,^40^ underscoring its potential to lastingly boost recovery though offline consolidation in rehabilitative settings.

From a functional stand point, the simultaneous combination of error- and reward-based learning might be more beneficial than each learning mechanism in isolation,^37^ as the reward signal might aid the motor system to interpret sensory prediction errors early after stroke. This view aligns with findings of improved motor performance in stroke patients receiving informative task-specific feedback.^42–45^ Critically, a study by Stanton and colleagues monitoring feedback provided by physiotherapists found that purely motivational feedback was four times as frequent as informative feedback,^46^ underlining the importance of translating our present findings into clinical practice.

### Limitations and future directions

We here combined monetary incentives as used in previous studies^11,17^ with feedback in the form of smiling or frowning emojis.^47^ Hence, it remains unclear which form of reinforcement (monetary versus social) drove the observed effects, which should be disentangled by future studies. Moreover, one might argue that a 30-minute break between learning and retention might be too short to support meaningful retention. However, as this relatively short period of time was sufficient to observe significant retention, extended periods might yield even more pronounced effects.

A potential motivational effect of reward feedback on motor learning complementing its role as a teaching signal cannot be excluded.^48,49^ For example, a commonly held view in sports psychology is that feedback after good trials leads to better performance than feedback after bad trials.^49,50^ It is frequently argued that feedback increases perceived self-efficacy, which facilitates allocation of attentional resources,^51^ leads to better performance^50^ and is a predictor for successful motor learning.^52^ Conversely, constant negative feedback arising from unsuccessful attempts to use the paretic arm in concert with punishment feedback provided during the task may have detrimental motivational effects in patients similar to those observed in athletes.^49^ Hence, motivational effects might contribute to the dissociative effects of punishment and reward on motor adaptation learning observed here.

Finally, the neurophysiological underpinnings of the observed effects remain speculative given the lack of neural data. Future research should therefore focus on elucidating the neural substrates of error- and reinforcement-based learning after stroke and shed light on the mechanisms driving the beneficial effects of reward.

## Conclusion

We here established for the first time that acute stroke patients can successfully perform error-based motor adaptation learning with their paretic hand by replicating phenomena known from healthy participants, such as adaptation learning, savings, and motor retention. In contrast to previous findings in healthy controls and chronic patients, punishment led to significantly slower learning than reward feedback. Conversely, combining reward- and error-based learning reduced initial movement errors and, more importantly, enhanced offline consolidation and retention. Our findings suggest that beneficial reward effects were driven by a reduction in motor noise arising from its instructive value as a teaching signal, enabling performance-dependent reward to guide the motor system towards more optimal motor control policies. Our current findings hold seminal clinical implications as they outline a combination of motor adaptation training with reward incentives as a promising avenue for neurorehabilitative interventions, aiming to support the retention of novel motor control policies early after stroke.

## Methods

### Participants

Twenty-four acute stroke patients were recruited from the Stroke Unit of the Department of Neurology at the University Hospital Cologne (time since stroke: mean = 5.3 days, std = 5.0 days; age: mean = 62 years, std = 11 years; 12 male, 12 female; see Supplementary Figure 1 for a lesion overlay). Two patients dropped out before the study was finalized. Inclusion criteria were (i) first-ever ischemic stroke and (ii) uni-lateral impairment of hand or arm motor function. Exclusion criteria were (i) bi-hemispheric infarcts, (ii) cerebral hemorrhage, (iii) prior infarcts, (iv) other neurological diseases, (v) severe aphasia or neglect, and (vi) a severe level of motor impairment (e.g., hemiplegia) that rendered study participation impossible. Subjects provided informed written consent prior to participation. The ethics committee of the Medical Faculty of the University of Cologne approved the study conducted in accordance with the Declaration of Helsinki.

### Task design and data collection

Patients performed a visuomotor rotation task with their paretic arm using a portable joystick in a bedside manner. Each trial consisted of a simple center-out reaching movement. Once participants had positioned the joystick cursor in the center circle (which was facilitated by the help of the joystick’s internal spring), the outside target turned red, indicating to start the reaching movement. A movement was considered complete once the cursor had reached the outside target. Participants were instructed to move as precisely and swiftly as possible. Importantly, due to the varying levels of stroke-related impairment, we decided against imposing a time limit on the movement as performance times drastically varied between patients. While this task might appear rather simple at first glance, extensive piloting prior to data collection had shown that the complexity level was the achievable maximum for more severely affected patients in the acute stage.

All patients participated in three sessions. During task performance, patients were lying in bed with a computer screen positioned in front of them on a height-adjustable table. The view of their paretic arm and hand was obstructed by a curtain. The experiment was divided into a learning period and a retention period (Figure 1). The learning period included a baseline of 15 trials (no rotation) after which a systematic visuomotor rotation was introduced (60 trials, adaptation block 1), followed by a de-adaptation block (no rotation, 15 trials) and a subsequent second adaptation block with cursor rotation (60 trials, adaptation block 2). After a 30-minute break, retention was assessed with 50 rotation trials, followed by a washout block of 25 trials (no rotation). The deviation angle used to induce the need for adaptation varied between 30, 40, and 50 degrees, depending on the session, to minimize carry-over effects from one session to another. The order of deviation angles across sessions was counter-balanced across participants. Of note, all adaptation blocks within a single session used the same rotation angle.

Performance was operationalized as *movement error*, quantified as the deviation from the optimal trajectory,^21^ i.e., the direct line from the center to the target. The movement error was computed via the cumulative distances between a subject’s reaching trajectory in a given trial and the optimal trajectory (see Supplementary Figure 2 for a schematic explanation). Thus, the movement error was influenced by both the accuracy of the movement (with more accurate movements resulting in smaller distances) as well as the speed of execution (with faster movements resulting in fewer sampled time points). In other words, faster and more precise movements resulted in a smaller movement errors and ideal behavior would have resulted in a movement error of zero.

In contrast to the majority of previous motor adaptation studies, we quantified performance using the movement error rather than the angular error at a specific point along the movement trajectory for several reasons specific to our sample of acute stroke patients. Extensive pilot testing showed that acute patients heavily relied on continuous online corrections of ongoing movements rather than changing their initial movement direction. Moreover, many acute patients were simply not able to initiate or perform smooth and continuous movements but produced jerkier movement trajectories instead. Such trajectories rendered a reliable detection of a meaningful angular error almost impossible (see Supplementary Figure 2).

During the learning phase of sessions 1 and 3, patients received either a performance-dependent monetary reward accompanied by a smiling emoji or a monetary punishment combined with a frowning emoji. Stimuli were taken from the Lisbon Emoji and Emoticon Database (LEED) to match arousal levels across conditions.^47^ In the reward condition, patients started out with a value of 0 Euros and gained money if their performance in a given trial exceeded the median performance of the past five trials.^53^ Conversely, participants started out with 30 Euros in the punishment condition and lost money if their performance in a given trial was worse than the median of the previous five trials. We chose a relative rather than an absolute reward/punishment criterion to account for the considerable amount of inter-individual variance in task performance across patients arising from varying levels of stroke-induced motor impairment. While the order of the reward and punishment sessions was counter-balanced across patients, session 2 served as a neutral control condition without reinforcement feedback in all patients. Notably, across all sessions, the retention period was carried out without reinforcement, as we aimed to probe retention effects in the absence of reinforcement feedback. This is particularly important as patients leaving a therapeutic setting have to retain a newly learned motor control policy without continuous performance feedback.

### Statistics

We calculated model-free and model-based analyses to quantify motor adaptation and retention. Model-free analyses compared motor adaptation performance operationalized as movement errors averaged across trials. To ensure normal distribution of data, the decimal logarithm of movement error data was used for model-free analyses. To avoid values below 1 becoming negative, 1 was added to the movement errors before the logarithmic transformation. For model-based analyses, we relied on a widely used single-rate state-space model to estimate learning rates for each block.^11^ This model predicts the future state 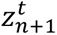 based on the equation

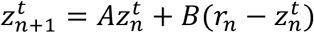

by combining the estimation of the current state 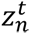 weighted by a retention rate A (also called decay rate or memory rate) with the difference between predicted and actual movement 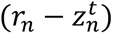 weighted by a learning rate B. We fitted the model separately for adaptation block 1, adaptation block 2, and the retention block. Data were filtered before model fitting to remove outliers and improve model fit. First, we created epochs based on the median of three consecutive trials. Next, we removed extreme outliers, i.e., trials exceeding the error observed in the first epoch of a block. We then applied a Hampel filter to each block leaving out the first two epochs per block. The derived learning rates were used for statistical comparisons.

### Effects of motor adaptation

First, we tested whether patients were able to exhibit motor adaptation by comparing the mean of the first 15 trials with the mean of the last 15 trials of adaptation block 1 by collapsing the data across all reinforcement conditions using a two-sided paired-samples t-test. We then tested for savings, i.e., whether adaptation was achieved faster when being introduced to the same rotation angle for a second time in a model-free and model-based way.^9^ For model-free analyses, we compared the mean of the first 15 trials of adaptation blocks 1 and 2. We additionally probed for savings in a model-based fashion by comparing the learning rates in adaptation blocks 1 and 2 using a two-sided paired sample t-test. To assess whether faster learning in the early learning phase might result in superior retention, we tested for a Pearson correlation between learning rates in adaptation block 1 and average movement errors in the retention block.

### Reinforcement effects

For model-free analyses, we tested for reinforcement effects on movement errors across blocks using a repeated measures ANOVA with the within-subject factor reinforcement condition for adaptation 1, adaptation 2, retention, and washout blocks. Here, each subject’s average movement errors per block were used as dependent variables. For the model-based approach, blockwise learning rates were compared between conditions. For all ANOVAs, a Greenhouse-Geisser correction was applied where appropriate. Post-hoc two-sided paired sample t-tests were FDR-corrected for multiple comparisons. To rule out that reinforcement had a general effect on overall motor performance rather than motor learning, we also tested for reinforcement differences in the baseline block using a repeated measures ANOVA.

After testing for blockwise reinforcement effects, we examined whether retention after a 30-minute break depended on the type of reinforcement received during the initial learning phase. To do so, we computed two-sided paired-samples t-tests for each condition, comparing the average movement error in adaptation block 2 with that in the retention block. Of note, successful retention can be reflected by either the absence of a significant difference or by an improvement in motor performance after the break.

### Speed-accuracy trade-off

To test whether actual learning occurred rather than a mere shift along the speed-accuracy continuum, we addressed both domains independently. Speed was operationalized as movement duration, i.e., the time it took participants to move the joystick from the center to the target. Accuracy was operationalized as movement inaccuracy, which was computed by dividing the movement error by the number of sampled points, thereby eliminating the influence of the time component. Again, the logarithm of the data (after adding 1 before the logarithmic transformation to avoid negative values) was used to ensure normal distribution. Using both measures, we repeated all aforementioned model-free analyses using movement duration or movement inaccuracy as the dependent variable.

### Motor noise

Motor noise was quantified as the standard deviation of movement inaccuracy. To test for the immediate, trial-wise effect of reinforcement feedback, we computed the standard deviation of the inaccuracy per subject across all trials (t_n+1_) occurring immediately after a trial that fulfilled the reward/punishment criteria (t_n_) within adaptation 1. These trials were compared to trials (t_n+1_) following trials without reinforcement (t_n_) using one-sided paired-sample t-tests. These analyses were conducted separately for the reward and punishment conditions. Of note, we expected a decrease in motor noise following rewarded trials in line with previous findings in healthy subjects.^12^ To test whether the effect of motor noise had a direct impact on movement execution, we repeated these analyses using average movement inaccuracy rather than motor noise as the outcome measure. We tested for a main effect of reinforcement on motor noise by using a repeated-measures ANOVA with the within-subject factor reinforcement condition for both the learning and the retention phase. To assess whether the level of motor noise influenced initial learning rates and retention, we tested for correlations of motor noise in adaptation 1 with learning rates during adaptation 1 and movement errors during the retention phase.

## Data availability

Data are available from the corresponding author upon reasonable request.

## Acknowledgements

This work was funded by the Deutsche Forschungsgemeinschaft (DFG, German Research Foundation) – Project-ID 431549029 – SFB 1451.

## Competing interests

The authors report no competing interests.

## Contributions

LJV, TP, GRF, and STG contributed to the conception and design of the study. TP, JGN, and VMW contributed to data collection. TP, FML, and LJV performed data analyses. TP, VMW, JGN, and LJV contributed to the design of the figures. TP and LJV drafted the manuscript. All authors contributed to the editing and revision of the manuscript.

## Supplementary Figures

**Supplementary Figure 1:**
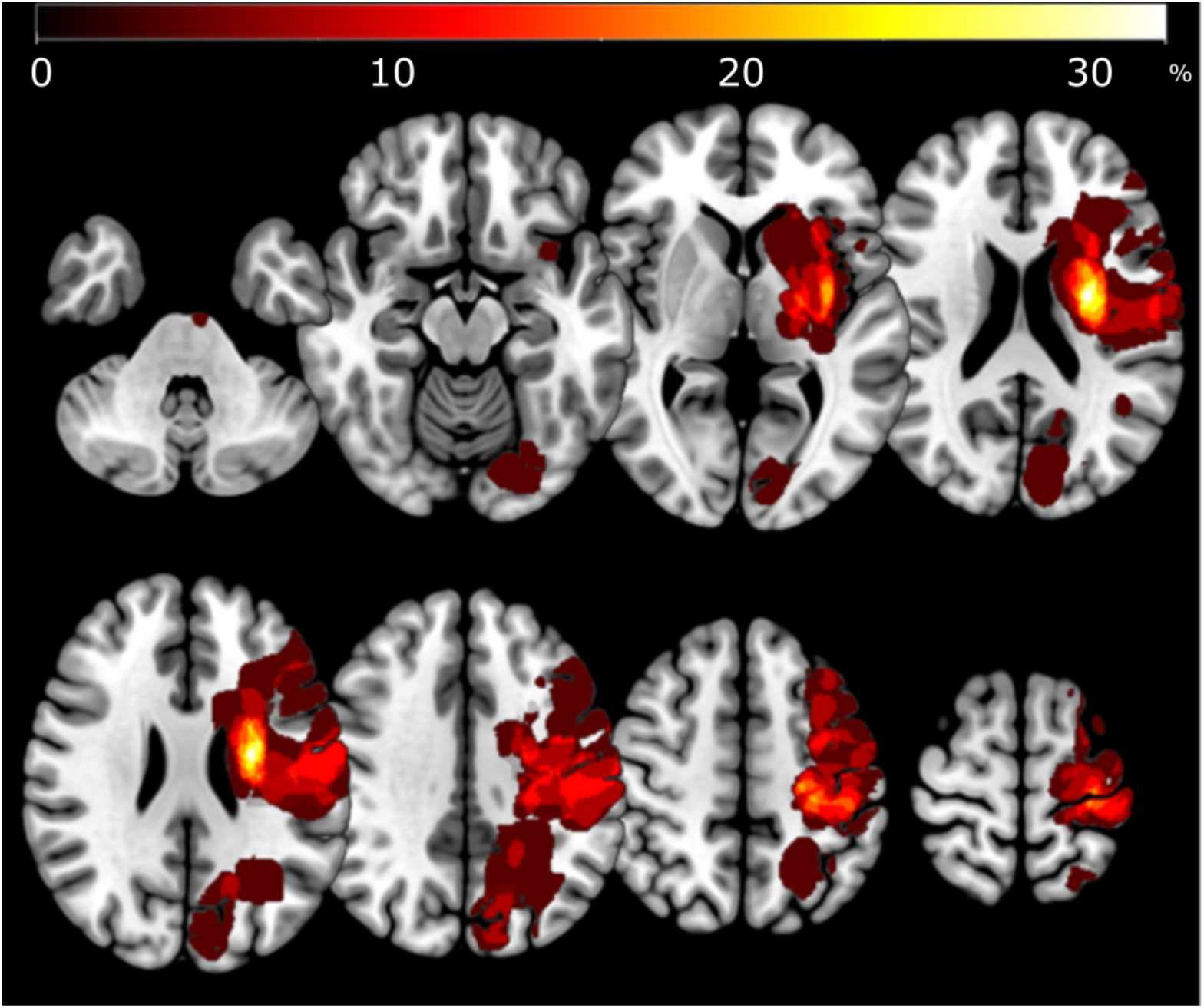
Lesion overlay. Please note that stroke lesions of all patients are projected on the same hemisphere for visualization purposes.

**Supplementary Figure 2:**
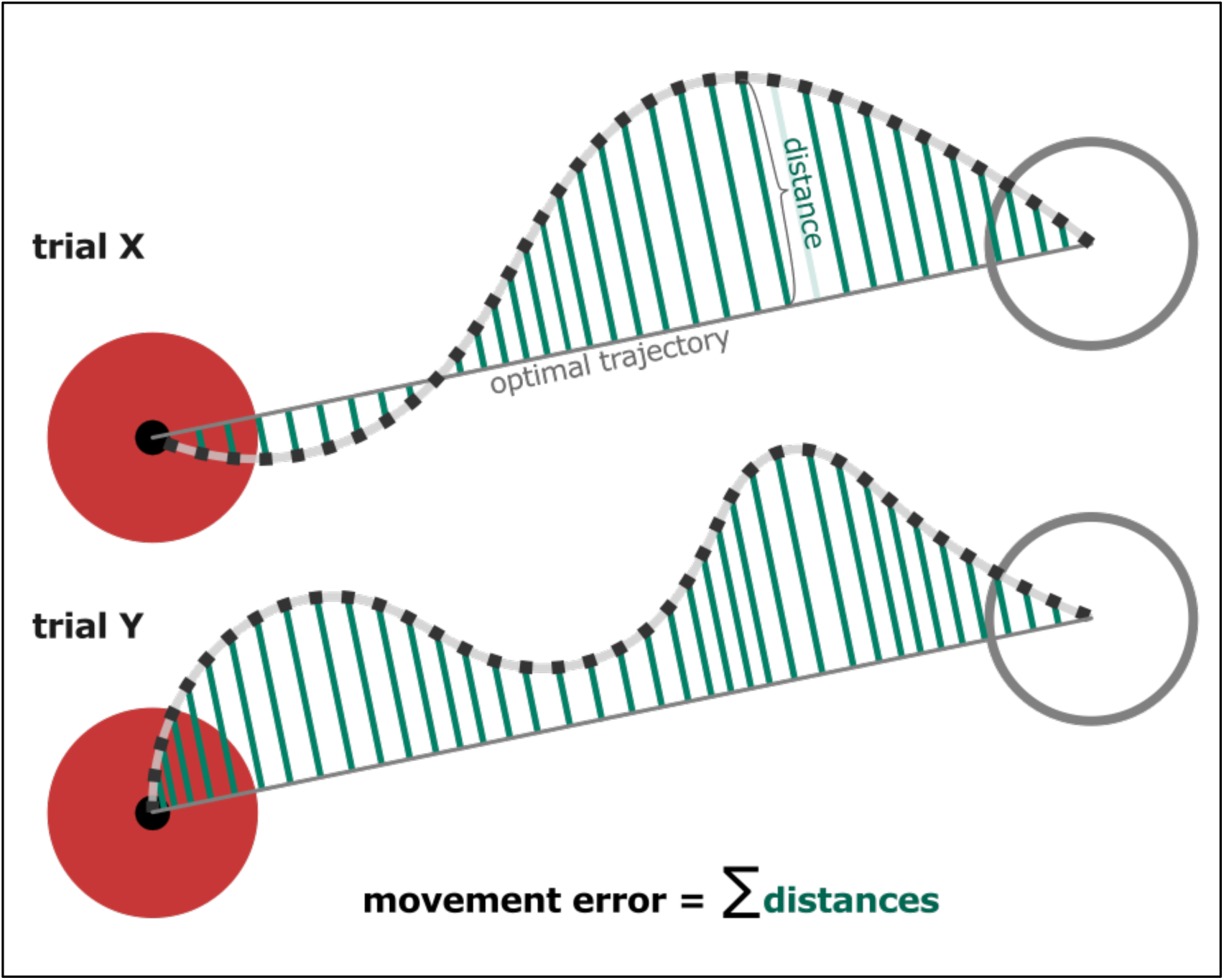
Schematic visualization of trial-wise movement error calculation. Movement errors were calculated as the sum of distances from the optimal trajectory, i.e., from the shortest possible line connecting the starting point with the center of the target. The Euclidian distances were computed between each sampled time point along the actual movement trajectory and the optimal trajectory. As the movement error was determined by summing up all distances in a given trial, it reflected both speed and accuracy: A higher movement error could either stem from a less accurate movement resulting in longer distances, or from a slower movement resulting in a higher number of sampled time points and hence a higher number of distances per trial.

## Notes

### Competing Interest Statement

The authors have declared no competing interest.

### Author Declarations

The ethics committee of the Medical Faculty of the University of Cologne gave ethical approval for this work.

### Summary of Updates

Additional analyses on the role of motor noise were included, allowing for a mechanistic interpretation of the beneficial effect of reward versus the detrimental effect of punishment. Moreover, we disentangled the contribution of changes in speed and accuracy, showing that patients exhibited actual learning rather than a mere shift along the speed-accuracy trade-off.

## References

1. Dobkin, B. H. Rehabilitation after Stroke. New England Journal of Medicine 352, 1677–1684 (2005).

2. Feigin, V. L. et al. Global, regional, and national burden of stroke and its risk factors, 1990–2019: a systematic analysis for the Global Burden of Disease Study 2019. Lancet Neurol 20, 795–820 (2021).

3. Krakauer, J. W. The Applicability of Motor Learning to Neurorehabilitation. Oxford Textbook of Neurorehabilitation vol. 1 (Oxford University Press, 2015).

4. Gregor, S., Saumur, T. M., Crosby, L. D., Powers, J. & Patterson, K. K. Study Paradigms and Principles Investigated in Motor Learning Research After Stroke: A Scoping Review. Arch Rehabil Res Clin Transl 3, 100111 (2021).

5. Roemmich, R. T. & Bastian, A. J. Closing the Loop: From Motor Neuroscience to Neurorehabilitation. Annu Rev Neurosci 41, 415–429 (2018).

6. Cashaback, J. G. A., McGregor, H. R., Mohatarem, A. & Gribble, P. L. Dissociating error-based and reinforcement-based loss functions during sensorimotor learning. PLoS Comput Biol 13, e1005623 (2017).

7. Izawa, J. & Shadmehr, R. Learning from Sensory and Reward Prediction Errors during Motor Adaptation. PLoS Comput Biol 7, e1002012 (2011).

8. Abram, S. J., Tsay, J. S., Yosef, H., Reisman, D. S. & Kim, H. E. The Detrimental Effect of Stroke on Motor Adaptation. Neurorehabil Neural Repair 39, 213–225 (2025).

9. Wolpert, D. M., Diedrichsen, J. & Flanagan, J. R. Principles of sensorimotor learning. Nat Rev Neurosci 12, 739–751 (2011).

10. Shadmehr, R., Smith, M. A. & Krakauer, J. W. Error correction, sensory prediction, and adaptation in motor control. Annu Rev Neurosci 33, 89–108 (2010).

11. Galea, J. M., Mallia, E., Rothwell, J. & Diedrichsen, J. The dissociable effects of punishment and reward on motor learning. Nat Neurosci 18, 597–602 (2015).

12. Manohar, S. G. et al. Reward Pays the Cost of Noise Reduction in Motor and Cognitive Control. Current Biology 25, 1707–1716 (2015).

13. Bonnì, S. et al. Intermittent Cerebellar Theta Burst Stimulation Improves Visuo-motor Learning in Stroke Patients: a Pilot Study. Cerebellum 19, 739–743 (2020).

14. Backhaus, W., Braass, H., Gerloff, C. & Hummel, F. C. Can Daytime Napping Assist the Process of Skills Acquisition After Stroke? Front Neurol 9, (2018).

15. Moore, R. T., Piitz, M. A., Singh, N., Dukelow, S. P. & Cluff, T. Assessing Impairments in Visuomotor Adaptation After Stroke. Neurorehabil Neural Repair 36, 415–425 (2022).

16. Scheidt, R. A. & Stoeckmann, T. Reach Adaptation and Final Position Control Amid Environmental Uncertainty After Stroke. J Neurophysiol 97, 2824–2836 (2007).

17. Quattrocchi, G., Greenwood, R., Rothwell, J. C., Galea, J. M. & Bestmann, S. Reward and punishment enhance motor adaptation in stroke. J Neurol Neurosurg Psychiatry 88, 730–736 (2017).

18. Koch, E. T., Dukelow, S. P. & Cluff, T. Motor learning after stroke: what we’ve learned and what lies ahead. Brain (2025) doi:10.1093/brain/awaf388.

19. Langhorne, P., Bernhardt, J. & Kwakkel, G. Stroke rehabilitation. The Lancet 377, 1693–1702 (2011).

20. Yin, C., Li, B. & Gao, T. Differential effects of reward and punishment on reinforcement-based motor learning and generalization. J Neurophysiol 130, 1150–1161 (2023).

21. Doost, M. Y. et al. Learning a Bimanual Cooperative Skill in Chronic Stroke Under Noninvasive Brain Stimulation: A Randomized Controlled Trial. Neurorehabil Neural Repair 33, 486–498 (2019).

22. Wu, H. G., Miyamoto, Y. R., Castro, L. N. G., Ölveczky, B. P. & Smith, M. A. Temporal structure of motor variability is dynamically regulated and predicts motor learning ability. Nat Neurosci 17, 312–321 (2014).

23. Baguma, M. et al. Preserved motor skill learning in acute stroke patients. Acta Neurol Belg 120, 365–374 (2020).

24. Haith, A. M., Huberdeau, D. M. & Krakauer, J. W. The influence of movement preparation time on the expression of visuomotor learning and savings. Journal of Neuroscience 35, 5109–5117 (2015).

25. Yin, C. & Wei, K. Savings in sensorimotor adaptation without an explicit strategy. J Neurophysiol 123, 1180–1192 (2020).

26. Huberdeau, D. M., Haith, A. M. & Krakauer, J. W. Formation of a long-term memory for visuomotor adaptation following only a few trials of practice. J Neurophysiol 114, 969–977 (2015).

27. Galea, J. M., Vazquez, A., Pasricha, N., Orban de Xivry, J.-J. & Celnik, P. Dissociating the Roles of the Cerebellum and Motor Cortex during Adaptive Learning: The Motor Cortex Retains What the Cerebellum Learns. Cerebral Cortex 21, 1761–1770 (2011).

28. Tseng, Y., Diedrichsen, J., Krakauer, J. W., Shadmehr, R. & Bastian, A. J. Sensory Prediction Errors Drive Cerebellum-Dependent Adaptation of Reaching. J Neurophysiol 98, 54–62 (2007).

29. Mazzoni, P. & Krakauer, J. W. An implicit plan overrides an explicit strategy during visuomotor adaptation. Journal of Neuroscience 26, 3642–3645 (2006).

30. Izawa, J., Criscimagna-Hemminger, S. E. & Shadmehr, R. Cerebellar contributions to reach adaptation and learning sensory consequences of action. Journal of Neuroscience 32, 4230–4239 (2012).

31. Albert, S. T. & Shadmehr, R. The Neural Feedback Response to Error As a Teaching Signal for the Motor Learning System. The Journal of Neuroscience 36, 4832–4845 (2016).

32. Sternad, D., Abe, M. O., Hu, X. & Müller, H. Neuromotor Noise, Error Tolerance and Velocity-Dependent Costs in Skilled Performance. PLoS Comput Biol 7, e1002159 (2011).

33. Thorp, E. B., Kording, K. P. & Mussa-Ivaldi, F. A. Using noise to shape motor learning. J Neurophysiol 117, 728–737 (2017).

34. Dhawale, A. K., Smith, M. A. & Ölveczky, B. P. The Role of Variability in Motor Learning. Annu Rev Neurosci 40, 479–498 (2017).

35. Lyons, E. M., Goldsworthy, M. R. & Hordacre, B. Exploring neural variability, movement variability and motor learning in healthy adults and people with stroke. Clinical Neurophysiology 175, 2110759 (2025).

36. Therrien, A. S., Wolpert, D. M. & Bastian, A. J. Effective reinforcement learning following cerebellar damage requires a balance between exploration and motor noise. Brain 139, 101–114 (2016).

37. Branscheidt, M. et al. Reinforcement Learning is Impaired in the Sub-acute Post-stroke Period. Neurorehabil Neural Repair 39, 297–311 (2025).

38. Nikooyan, A. A. & Ahmed, A. A. Reward feedback accelerates motor learning. J Neurophysiol 113, 633–646 (2015).

39. Widmer, M., Ziegler, N., Held, J., Luft, A. & Lutz, K. Rewarding feedback promotes motor skill consolidation via striatal activity. Prog Brain Res 229, 303–323 (2016).

40. Abe, M. et al. Reward Improves Long-Term Retention of a Motor Memory through Induction of Offline Memory Gains. Current Biology 21, 557–562 (2011).

41. Vassiliadis, P. et al. Reward boosts reinforcement-based motor learning. iScience 24, 102821 (2021).

42. Rendos, N. K. et al. Verbal feedback enhances motor learning during post-stroke gait retraining. Top Stroke Rehabil 28, 362–377 (2021).

43. van Vliet, P. M. & Wulf, G. Extrinsic feedback for motor learning after stroke: What is the evidence? Disabil Rehabil 28, 831–840 (2006).

44. Subramanian, S. K., Massie, C. L., Malcolm, M. P. & Levin, M. F. Does Provision of Extrinsic Feedback Result in Improved Motor Learning in the Upper Limb Poststroke? A Systematic Review of the Evidence. Neurorehabil Neural Repair 24, 113–124 (2010).

45. Cirstea, M. C. & Levin, M. F. Improvement of Arm Movement Patterns and Endpoint Control Depends on Type of Feedback During Practice in Stroke Survivors. Neurorehabil Neural Repair 21, 398–411 (2007).

46. Stanton, R., Ada, L., Dean, C. M. & Preston, E. Feedback Received While Practicing Everyday Activities During Rehabilitation After Stroke: An Observational Study. Physiotherapy Research International 20, 166–173 (2015).

47. Rodrigues, D., Prada, M., Gaspar, R., Garrido, M. V. & Lopes, D. Lisbon Emoji and Emoticon Database (LEED): Norms for emoji and emoticons in seven evaluative dimensions. Behav Res Methods 50, 392–405 (2018).

48. Chiviacowsky, S. The motivational role of feedback in motor learning. in Advancements in Mental Skills Training 44–56 (Routledge, 2020). doi:10.4324/9780429025112-5.

49. Lewthwaite, R. & Wulf, G. Optimizing motivation and attention for motor performance and learning. Curr Opin Psychol 16, 38–42 (2017).

50. Saemi, E., Porter, J. M., Ghotbi-Varzaneh, A., Zarghami, M. & Maleki, F. Knowledge of results after relatively good trials enhances self-efficacy and motor learning. Psychol Sport Exerc 13, 378–382 (2012).

51. Themanson, J. R. & Rosen, P. J. Examining the relationships between self-efficacy, task-relevant attentional control, and task performance: Evidence from event-related brain potentials. British Journal of Psychology 106, 253–271 (2015).

52. Chiviacowsky, S., Wulf, G. & Lewthwaite, R. Self-Controlled Learning: The Importance of Protecting Perceptions of Competence. Front Psychol 3, 1–8 (2012).

53. Widmer, M., Lutz, K. & Luft, A. R. Reduced striatal activation in response to rewarding motor performance feedback after stroke. Neuroimage Clin 24, 102036 (2019).

